# Low-Cost Enhancement of Facial Mask Filtration to Prevent Transmission of COVID-19

**DOI:** 10.1101/2020.08.01.20166637

**Authors:** Hari Bhimaraju, Ramesh Jain, Nitish Nag

## Abstract

The use of face masks is recommended worldwide to reduce the spread of COVID-19. A plethora of facial coverings and respirators, both commercial and homemade, pervade the market, but the true filtration capabilities of many homemade measures against the virus are unclear and continue to be unexplored. In this work, we compare the efficacy of the following masks in keeping out particulate matter below 2.5 microns: N95 respirators, surgical masks, cloth masks, cloth masks with activated carbon air filters, cloth masks with HVAC air filters, lightly starch-enhanced cloth masks, and heavily-starched cloth masks. The experiments utilize an inhalation system and aerosol chamber to simulate a masked individual respiring aerosolized air. COVID-19 disproportionately affects people in low-income communities, who often lack the resources to acquire appropriate personal protective equipment and tend to lack the flexibility to shelter in place due to their public-facing occupations. This work tests low-cost enhancements to homemade masks to assist these communities in making better masks to reduce viral transmission. Experimental results demonstrate that the filtration efficacy of cloth masks with either a light or heavy starch can approach the performance of much costlier masks. This discovery supports the idea of low-cost enhancements to reduce transmission and protect individuals from contracting COVID-19.

## I. Introduction

Coronavirus disease 2019 (COVID-19), also known as severe acute respiratory syndrome coronavirus 2 (SARS-CoV-2), is a highly infectious virus that has changed the modern global landscape with over 15 million confirmed cases and 633,000 deaths as of this paper submission and growing [20]. COVID-19 is transported primarily through respiratory droplets and contact routes. Respiratory droplets are droplet particles with a diameter between 5 *µ*m and 10 *µ*m, and contact routes include direct contact of mucosal surfaces with infected people or fomites [24]. A sneeze can generate as many as 40,000 droplets, which can evaporate to particles in the 0.5*µ*m to 12*µ*m range [6]. During this pandemic, numerous countries deemed the use of cloth masks in public mandatory or highly encouraged to minimize the ability of sneezes and other droplet-emitting actions to spread the virus [13]. The CDC released a statement supporting the efficacy of masks in reducing the spread of the virus from infected individuals with and without external symptoms [4]. However, healthcare personal protective equipment (PPE) resources are already stretched thin, and encouraging the public to purchase masks is further dwindling hospitals’ limited supplies [2]. Low-income communities suffer disproportionately from COVID-19, having both a higher risk for transmission and a lower probability of recovering fully. A large factors of this is due to the lack of PPE [11]. To combat these issues, people make cloth masks at home, both for personal use and to donate to communities in need. Although these efforts are with good intent, the efficacy of these cloth masks in providing adequate protection is unclear. A study before the COVID-19 pandemic shows that penetration of cloth masks by sodium chloride particles within a specified size range is almost 97% while that of medical masks is 44% [14]. For this reason, the ability to manufacture low-cost facial masks with higher performance at home is especially valuable in these low resource communities.

The availability of resources for both hospitals and civilians is increasingly limited. Thus, we study the potential of low-cost materials and homemade masks in reducing the spread of the virus to spare medical resources for hospitals and encourage the use of effective filtration techniques in everyday life. In this work, we test various low-cost mask systems alongside popular medical and high-performance masks such as N95 masks. N95 masks use electrostatic filtration to trap particles [1]. Sars-CoV2 has amino acids that are chemically attached to sugars which can be charged, and is negatively charged at a neutral pH [1]. Thus, electrostatic surfaces like that of an N95 mask trap virus particles very effectively, adding an extra layer of filtration beyond the physical barriers of a mask [5]. To analyze the efficacy of the various masks in reducing the spread of the virus, we measure their ability to block aerosol particles that are similar to respiratory droplets that transmit COVID-19.

### II. Mask Types

We test a variety of masks which are described below.

### A. N95

N95 respirators are Filtering Facepiece Respirators, FFRs, that filter at least 95% of airborne particles. They are designed to seal tightly around the user’s mouth and nose, although some leakage is still expected. N95 respirators currently must be disposed of, after each use. They limit the user’s exposure to particles like the small aerosol droplets in which the virus travels, but they do not necessarily filter the air that the user exhales [9]. We use a variety of N95 mask models and brands, including some with extra filtration; this accounts for the variation of data observed within N95 results.

### B. Surgical

Surgical masks are face masks that are meant to block large-particle contaminants such as sprays or splatters from reaching the user’s mouth or nose [19]. They are designed to be loose-fitting and do not protect from small airborne particles, such as those disseminated from coughs or sneezes [19]. Surgical masks are single-use, disposable face coverings.

### C. Cloth

Cloth masks are face coverings that are widely available for the public and can be easily made at home. The CDC recommends that people use cloth masks whenever they exit their homes, because they may prevent virus-carrying individuals from spreading it to others [4] They vary in design and material, but in this study, we use a wide range of cotton fabrics to best mimic usage in the public [15]. We also include cloth masks made from the government-supplied mask fabric distributed in the slums of Tirupati, Andhra Pradesh, India.

### D. Activated Carbon Air Filter

Activated carbon, or activated charcoal, air filters use adsorption to purify the air of volatile organic compounds, odors, and other pollutants [18]. We use the cloth mask design with a pocket for the activated carbon filter.

### E. HVAC Air Filter

HVAC air filters are commonly used in heating, vacuuming, and air conditioning to filter a variety of pollutants and biological contaminants [8]. HVAC filters are effective in filtering particles as small as 0.3 microns [8]. We place an HVAC filter in the cloth mask pocket. We use a variety of cloth masks, which explains the variability in the performance of the HVAC filter masks.

### F. Starch

Rice is a staple food in many Asian communities and is consumed around the world [7]. Rice starch can be easily extracted by draining boiling rice before it is fully cooked. Starch is commonly used to stiffen cloth. We make starched cloth masks to better filter and trap aerosol particles that can potentially carry the virus. Starch has two components: amylose and amylopectin. The former has a linear structure of glucose molecules, while the latter has a branching structure. Long-grain rice varieties have higher amylose content while short-grain rice varieties have higher amylopectin content [17]. Due to its structure and hydroxyl groups, amylopectin is easily dissolved in water, so longer grain rice varieties produce less starch because of their higher swelling temperatures [23]. The government-supplied rice for the slums of Tirupati, Andhra Pradesh, India is medium-grain, so we use the same type to accurately replicate their amylose and amylopectin content. The hydroxyl groups in starch make starch a polar molecule. This may give starched cloth masks both the physical and polar barrier capabilities of an N95 mask [16]. The procedure for producing the starch masks is as follows: (1) cook rice in a 1:5 rice-to-water ratio, (2) sieve out the starchy water when the rice is 75% cooked, (3) soak a cloth mask overnight in the starch water. For the light starch mask, we squeeze out the starchy water from the mask in the morning and allow it to dry. For the heavy starch mask, we allow the mask to dry naturally without squeezing out the water. Although the heavy starch mask is more effective in filtering out particulate matter, the light starch mask is less stiff and offers more comfort.

### G. Price Comparison

The price of masks can be a key concern in low-income communities. We look through CDC-recommended mask models, average mask prices in various online providers, and speak to community groups overseeing mask making in rural India to identify accurate prices. Many masks and materials, such as N95 respirators, surgical masks, activated carbon filters and masks, and HVAC air filters, are typically sold in bulk. We divide the bulk prices by the number of mask units to display the per-unit prices of masks and materials in Figure 1. Activated Carbon (filter only) refers to the per-unit price of purchasing an Activated Carbon filter in a pack, while Activated Carbon Cloth Mask is the price of purchasing a pre-made mask. Cloth (low-income India) masks are charity-distributed masks in the city of Tirupati, Andhra Pradesh, India [21]. Cloth (retail USA) masks are the masks found when one searches for cloth masks on Amazon or Google in the US.

**Fig. 1.**
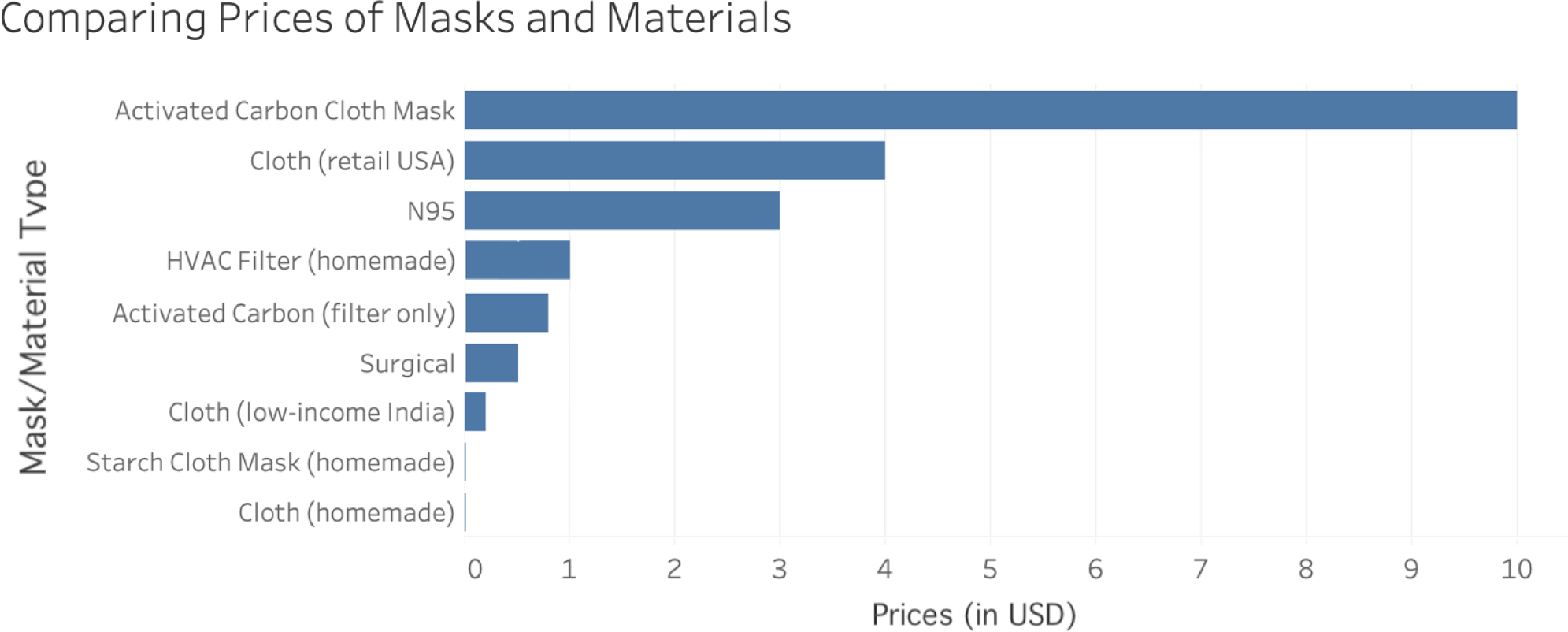
Comparing per-unit mask and material prices. This graph shows per-unit mask prices, either sold individually or sold in packs. Activated Carbon (filter only) refers to the per-unit price of purchasing an Activated Carbon filter in a pack, while Activated Carbon Cloth Mask is the price of purchasing a pre-made mask with the filter. Cloth (low-income) refers to the price of charity-distributed masks in rural India, while cloth (retail USA) masks are masks that are typically sold online in the USA. The most affordable masks for low-income communities are homemade cloth masks, starched cloth masks, and surgical masks.

### III. Methods and Experimental Design

We simulate an individual breathing air containing respiratory droplets through a mask by building a chamber in which a masked mannequin inhales the aerosolized 0.9% saline solution, a polar molecule. N95 masks serve as the gold standard and no masks serve as a negative control. Experiments record the efficacy of surgical masks, cloth masks, cloth masks with activated carbon air filters, cloth masks with HVAC air-filters, lightly starched cloth masks, and heavily starched cloth masks in comparison to the controls. The experiments collect particulate matter readings through the use of no mask, N95 mask, surgical mask, activated charcoal carbon cloth mask, air conditioning filter cloth mask, light starch cloth mask, heavy starch cloth mask, and regular cloth mask in an aerosolized setting. Our design for the cloth masks, shown in Figure 2, is worn like an N95 mask and tied in the back to maximize facial coverage and minimize leakage. Figure 3 shows the pores of a starched cloth mask beside the pores of a regular cloth mask to highlight starch’s ability to reduce pore size in cloth masks.

**Fig. 2.**
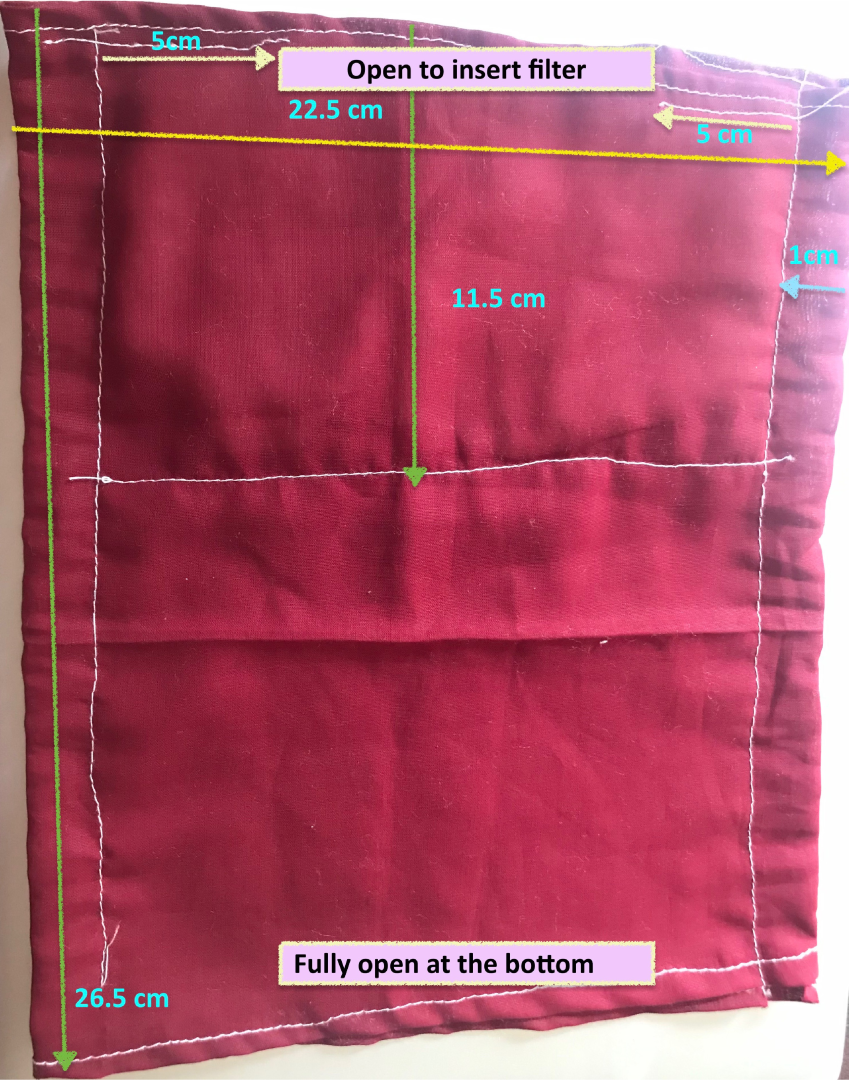
Cloth mask design. Folded measurements and a pocket for an additional filter, such as the Activated Carbon or HVAC filters are shown. The open part of the mask folds under the mannequin’s chin, covering the entire region from the mannequin’s nose to its neck.

**Fig. 3.**
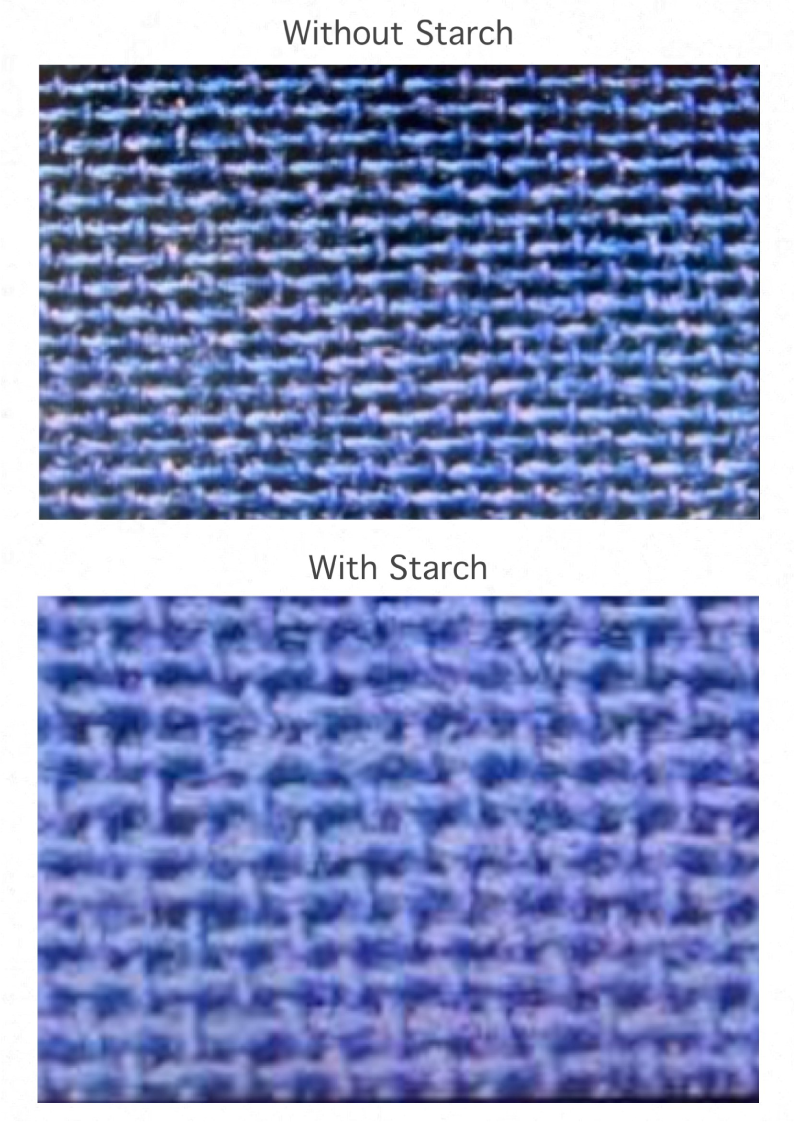
Microscope images at 200x magnification of regular cloth mask pores vs. starched cloth mask pores. The latter pores are visibly clogged with starch, allowing the mask to be more effective in filtering aerosol droplets.

We compare the PM2.5 concentration of each mask across numerous experimental trials, using no mask data as a negative control and N95 mask data as a positive control. These values represent how many aerosol particles of size 2.5 microns or less pass through the mask. Although respiratory droplets typically are in the 5 to 10-micron size range, droplet nuclei or aerosols in the size range of 0.3 to 5 microns can travel farther and remain in the air for longer [22] [10]. In this paper, particles of size 2.5 microns and below (PM2.5) remain the primary focus to conservatively account for droplet nuclei, considering the assumption that masks effective at filtering such minuscule particles will also be effective in filtering larger droplets.

The materials used in this experiment include the masks, a 30 L plastic container with an airtight lid, a Styrofoam mannequin head, two PMS5003 dust laser sensors from the Atmosome Measurement System, two 12000 RPM mini DC motors with attached plastic propeller blades, two L293D motor drive shields, an Arduino Uno, one 2 cm x 24 cm vacuum tube, an aerosol nebulizer, and saline solution [3]. Figure 4 displays the experimental setup.

**Fig. 4.**
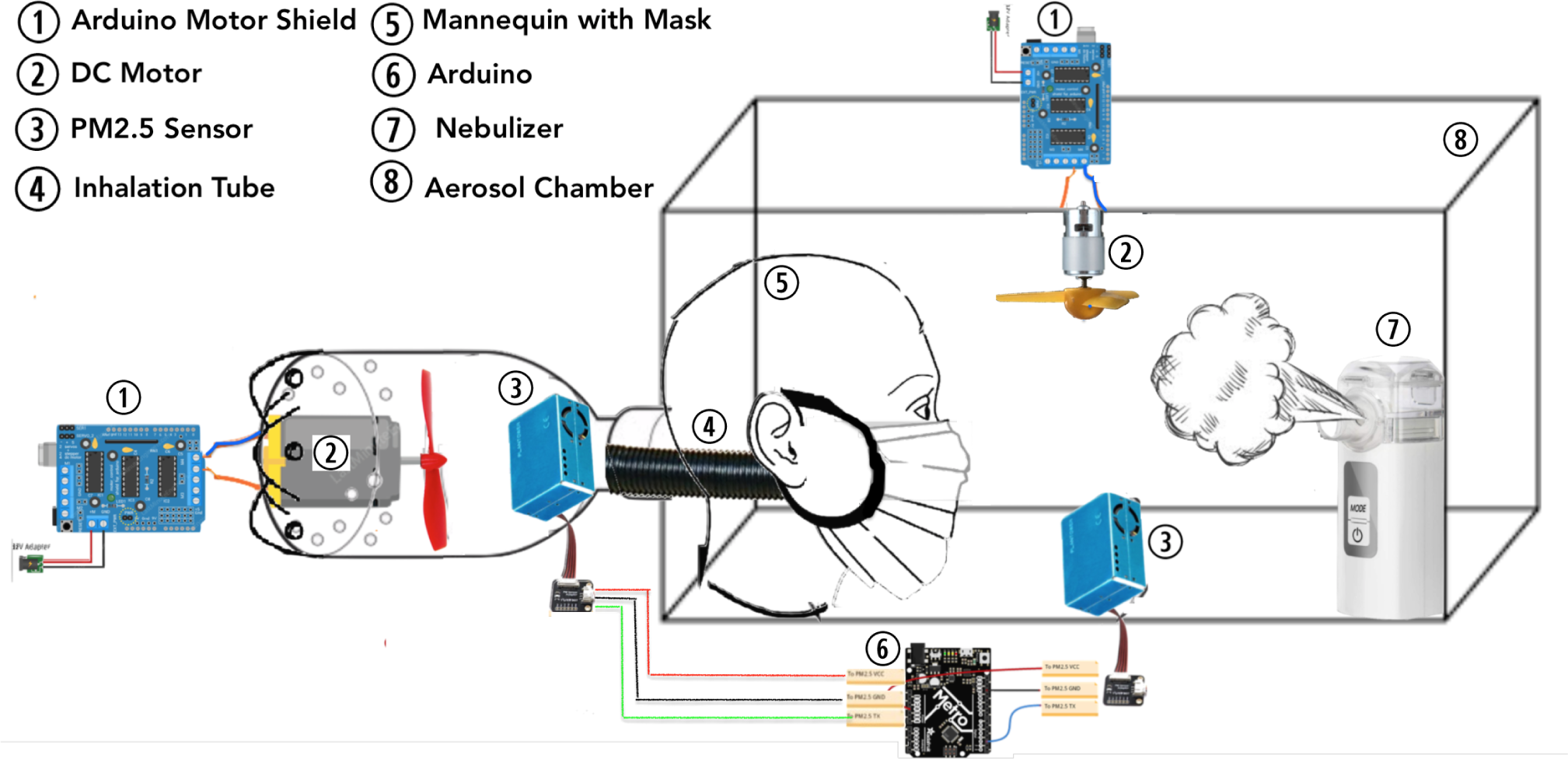
Experimental setup. Primary components include the inhalation simulation (left half of setup until end of mannequin) and aerosol chamber design (rectangular prism with sensor and fan). The materials in this setup include the masks, a 30 L plastic container with an airtight lid, a Styrofoam mannequin head, two PMS5003 dust laser sensors, two 12000 RPM mini DC motors with attached plastic propeller blades, two L293D motor drive shields, an Arduino Uno, one 2 cm x 24 cm vacuum tube, an aerosol nebulizer, and saline solution.

### A. Aerosol Particle Generation

A MAYLUCK Handheld Mesh Atomizer Nebulizer, a device that converts liquid into tiny aerosol particles, simulating sneeze or cough droplets. When the nebulizer is switched on, a ceramic piezoelectric crystal in it vibrates emitting high energy ultrasonic waves that pass through the solution and emit tiny aerosol particles through a micromesh. Wound wash sterile saline solution, consisting of purified water and 0.9% sodium chloride, represents respiratory aerosols that the virus travels through. Since the saline solution is polar, it more accurately represents the chemical properties of a virus [1]. The aerosolized liquid then pops out through the micromesh holes of the metal in atomized particles.

### B. Aerosol Chamber

A plastic container with a foam-lined, airtight lid houses the mannequin and simulates an indoor room in which an individual could be resting that contains aerosols that the virus could travel in. The mannequin head sits at the end of the chamber, with its inhalation system resting outside the box through a hole. The aerosol nebulizer sits at the other end, with another PMS5003 sensor, DC motor fan. We regulate a constant level of PM2.5 particles in this chamber by ensuring that the PMS5003 sensor values remain within a constant range. The DC motor fan blows down directly onto the aerosol stream to ensure that the particles circulate throughout the chamber.

### C. Inhalation System

An Arduino-based suction pump system emulates respiration. A 12000 RPM Mini Magnetic Motor with propeller blades affixes to the inside bottom of a 9 cm diameter bottle. The bottom has two circles of five 0.5 cm holes each drilled. The mouth of the bottle is attached to a lightweight plastic vacuum tube, 2 cm diameter and 24 cm long, and sealed with hot glue. The motor is connected to a motor shield and Arduino outside the bottle and plugged into a power socket. We programmed the motor to mimic tidal breathing with cycles consisting of 2 seconds inhale and 3 seconds pause (exhale), totaling 12 breaths per minute like a human being at rest. As the purpose of the circuit was to test the filtration capabilities of a mask, the exhale was not done through the mask, but rather simulated by pausing the motor.

### D. Procedure

First, we turn on the aerosolizer and allow the sodium chloride particulate count of size 2.5 microns and less to build up to an average saturation range of 8500 to 9500. We maintain this concentration in the aerosol chamber throughout the trial. Next, the mannequin begins respiration through the mask. We begin recording data after 45 seconds, to allow for the sensor to stabilize its output. Then, we record PM2.5 readings from the mannequin’s sensor as the mannequin respires for 2 minutes. With a resting breathing rate of 12 breaths/minute, alternate breaths are recorded to allow for maximum variability in the data. Each mask type has 16 individual trials with new masks in each trial. Between each measurement, we ventilate the container to reset the particulate matter count. We cycle through all masks before conducting a second trial, again maximizing the variability of data between masks of the same type. Figure 5 displays this nested experimental design. The means of every experimental trial and the comprehensive data sets are important for analysis, because the former provides independent data points and the latter provides a larger pool of data. We analyze the results through raw data visualizations, ANOVA models, nonparametric tests, and post-hoc tests on both the mean subsets and the whole data. We clean, visualize, and analyze the data using several Python packages, including NumPy, Pandas, StatsModels, Pingouin, SciPy, and Matplotlib.

**Fig. 5.**
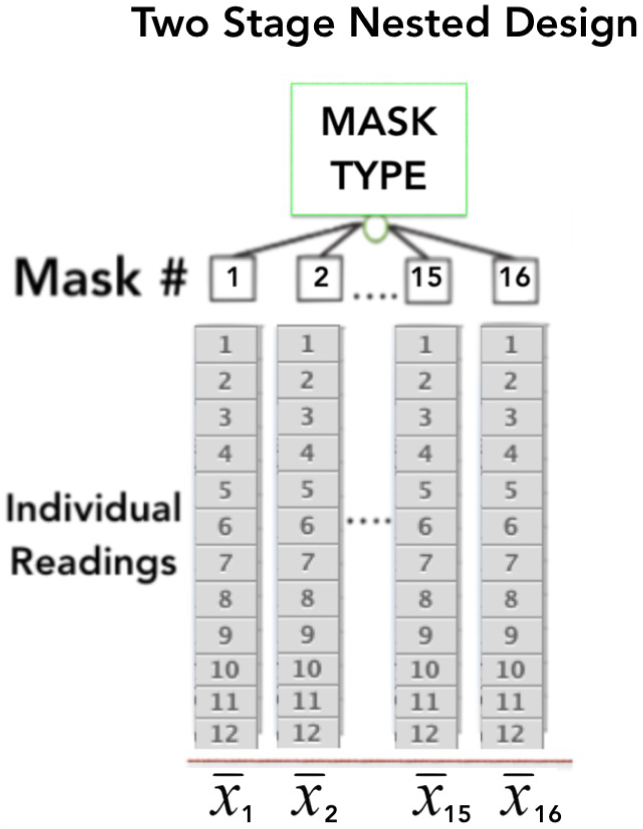
Nested experimental design. For each type of mask, we conduct 16 independent experimental trials with new mask models in each trial. Each trial consists of 12 individual readings. We study both the means of these trials and the comprehensive data in the experimental results.

## IV. Experimental Results

Results show that activated carbon masks and HVAC air filter masks are nearly as effective as N95 respirators. Surgical masks and heavy starch cloth masks are almost equally effective, followed closely by light starch cloth masks. Regular cloth masks do provide some filtration in comparison to the no mask control, but not by much. These findings can inform actionable public health and safety recommendations, because they suggest that low-cost enhancements to masks can be performed within households to significantly improve mask performance and, ultimately, reduce the spread of the virus.

Figure 6 shows the distribution of PM2.5 concentrations for each mask over a 2-minute series of readings. This graph suggests that the masks have varying filtration capabilities. Figure 7 visualizes the spread of PM2.5 concentration across each mask type.

**Fig. 6.**
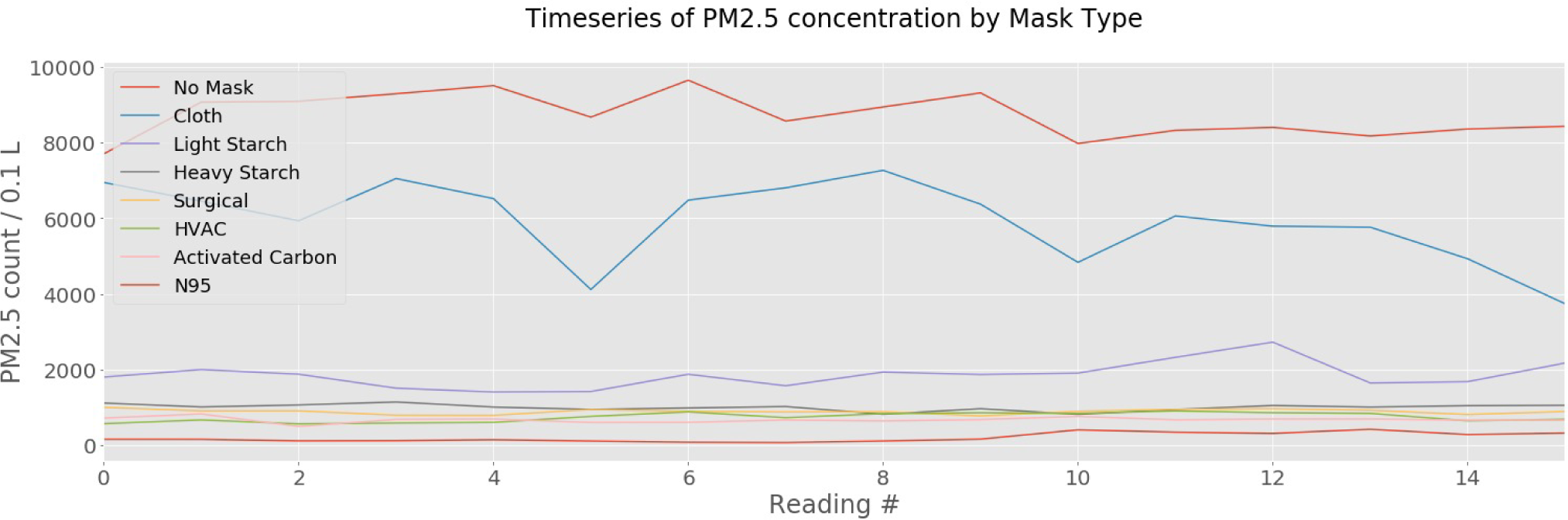
Time series of concentration of particulate matter below 2.5 microns (PM2.5) that passes through each mask over a 2-minute period during respiration. We record data at alternate breaths to allow for maximum variability. This plot suggests that the efficacy of the masks takes the following order, from most to least effective at filtration of PM2.5: N95 Respirator, HVAC filter and Activated Carbon filter, Surgical Mask and Heavy Starch Mask, Light Starch Mask, Cloth Mask.

**Fig. 7.**
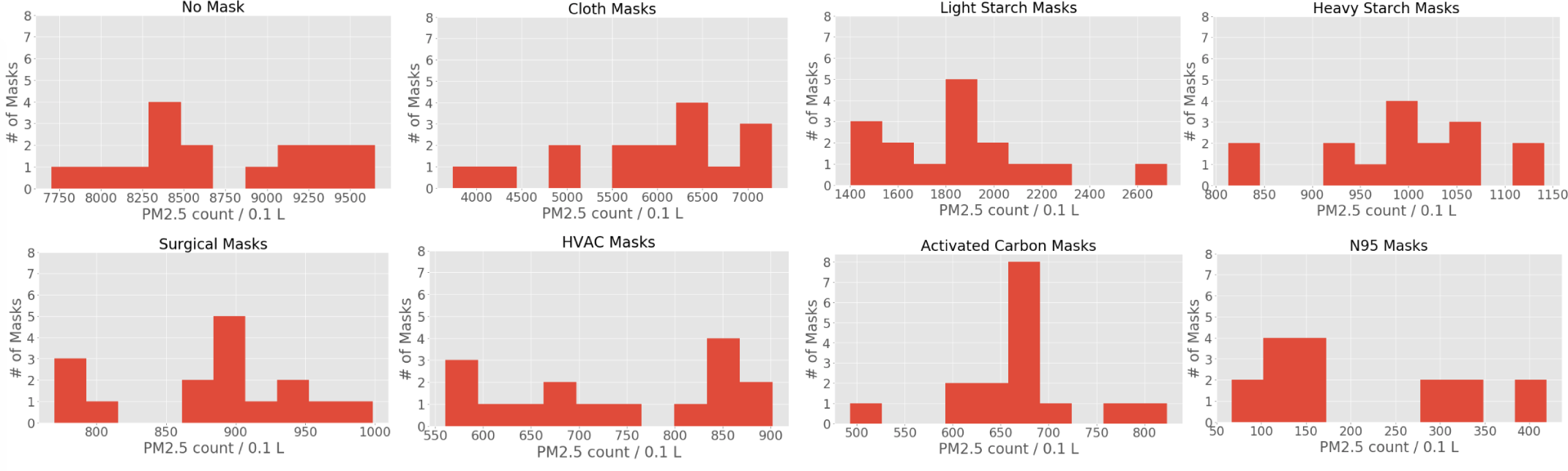
Distributions of the mean concentrations of particulate less than or equal to 2.5 microns that pass through each mask, in individual trials. The wider spread of data indicates a higher permeability of particulate matter in the cloth mask, with the distribution skewed toward higher particulate concentrations than the other masks. Some distributions look fairly normal, while others do not. The small sample sizes make it difficult to determine whether parametric tests, which require data to be in normal distributions, can be performed or not.

### A. Analysis of Means of Independent Trials

A box-and-whisker plot of the average PM2.5 concentration of across each of 16 trials for a given mask and the summary statistics for the masks, ordered from least to most effective, are displayed Figures 8 and Table I. The statistics indicate that the mean concentration of PM2.5 inhaled by the mannequin wearing a lightly starched cloth mask provides a three-fold improvementment in filtration ability over that of a regular cloth mask. The differences in box sizes in Figure 8 and the varying standard deviations across mask types in Table I reflect that the data lacks homogeneity of variance between categories; the Bartlett’s test for equality in variances confirms these observations. Thus, we use the Welch ANOVA model, as opposed to another model which assumes homogeneity of variance, to confirm the statistical significance of the results shown in Figures 6 and Figure 8.

**TABLE I.** Summary statistics of each type of mask, listed from least to most effective. Each row represents experimental trials with 16 individual masks of each type. The masks with higher mean concentrations, listed from top to bottom, have lower filtration capabilities and are less effective.

|  | # values | Mean | Std Dev | Min | 25% | 50% | 75% | Max |
| --- | --- | --- | --- | --- | --- | --- | --- | --- |
| No Mask | 16 | 8717.516 | 569.053 | 7702.583 | 8350.146 | 8621.167 | 9142.354 | 9650.166 |
| Cloth | 16 | 5940.385 | 1043.017 | 3738.333 | 5554.25 | 6215.333 | 6590.938 | 7266.667 |
| Light Starch | 16 | 1854.521 | 344.309 | 1402.417 | 1624.521 | 1869.292 | 1947.771 | 2722.25 |
| Heavy Starch | 16 | 996.984 | 89.789 | 812.833 | 957.667 | 1007.25 | 1048.458 | 1141 |
| Surgical | 16 | 884.5 | 66.134 | 769.833 | 857.104 | 891.708 | 924 | 998.25 |
| HVAC | 16 | 736.286 | 122.182 | 560.667 | 631.583 | 741.833 | 842.104 | 901.833 |
| Activated Carbon | 16 | 668.271 | 71.613 | 493.167 | 653.146 | 671.417 | 688.333 | 823.333 |
| N95 | 16 | 203.5 | 119.872 | 66.833 | 111.208 | 152.5 | 310.375 | 418.833 |

**Fig. 8.**
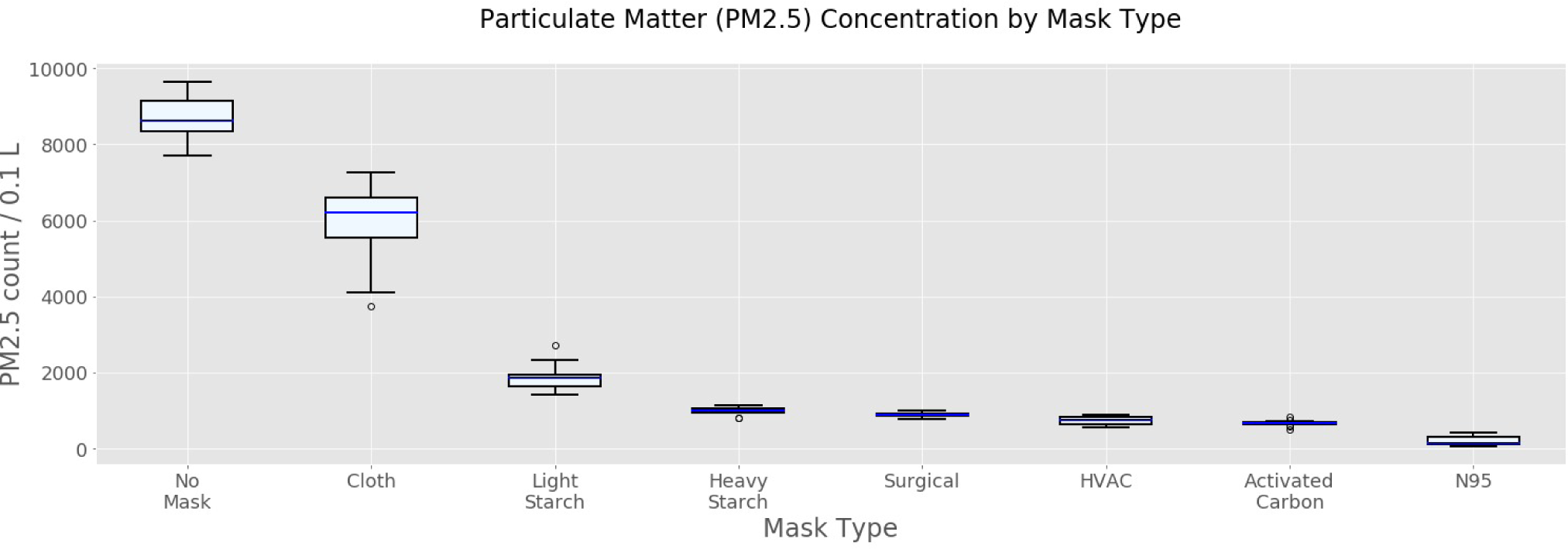
Box-and-whisker plot of average concentration of particulate matter below 2.5 microns (PM2.5) that passes through each mask during respiration in each experimental trial. This shows the quartiles of data from 10 independent experimental trials with new masks in each trial. This plot ranks the efficacy of the masks from most to least effective at filtration of PM2.5 based on mean filtration performance

We adopt a conservative significance standard, with a p-value ¡ 0.001 considered significant in our results. The Welch ANOVA test returns an F-statistic of 554.83 and a p-value much lower than 0.001, concluding that there is sufficient evidence for at least one of the mask types to be significantly different from the others in filtering particulate matter under 2.5 microns.

We examine these differences by performing pairwise t-tests between each mask type and the cloth mask, with the assumption of equal variance set to false. Since the p-values are far less than 0.001, there is sufficient evidence that each mask type is significantly different from the cloth mask. For instance, this shows evidence that the regular cloth mask allows more particulate matter to permeate through it, confirming that starching a cloth mask provides significantly better protection.

We check our assumptions for normality using the Shapiro-Wilks test and Q-Q plots. The independent trials for each mask type do reflect a normal distribution, but the small sample sizes make it difficult to assume normality despite these results. Non-parametric tests allow for a conservative verification of the previous parametric results because they do not assume normality.

We conduct the non-parametric Kruskal-Wallis test, which returns a statistic of 121.05 and a p-value much below 0.001. These results confirm again that the samples are not drawn from a similar distribution and that there is sufficient evidence that at least one of the mask types is significantly different from the others in filtering particulate matter.

As the Kruskal-Wallis test does not indicate which samples differ or by how much, we use the Mann-Whitney U test, a non-parametric statistical significance test for determining if two samples are drawn from the same distribution, to confirm the results of the t-tests. In this test, all the other types of masks compare against the cloth mask. All the tests yield the following result: the result statistic of 0.000 indicates that all the particulate values for the cloth sample are greater than all the values for the light-starch, heavy-starch, N95, activated carbon, HVAC filter, and surgical samples. The p-value is less than 0.001, indicating that the samples belong to different distributions.

### B. Analysis of the Comprehensive Data Set

In order not to violate the assumption of independence between observations in the aforementioned tests, the results only analyze the means of experimental trials instead of the entire data sample. In this section, we leverage the complete data set described in the nested experimental setup of Figure5. This significantly increases the sample size per mask trial from 1 mean to 12 readings, producing a resultant data set of 192 readings per mask type, as opposed to the 16 readings used in the Welch ANOVA. The parametric repeated measures ANOVA test is for related groups, such as those in the nested model, and does not rely on the independence of observations. The Greenhouse-Geisser corrected p-value adjusts for high sphericity, or unequal variance, in the repeated measures ANOVA test. The resulting p-value is much below 0.001, confirming that the statistical significance in the difference between each mask type is true for larger data sets as well.

Although the Shapiro-Wilks test shows some evidence that the complete data set is not normally distributed, this result is not fully conclusive due to the small sample size. We also implement the Friedman Test, the non-parametric version of the repeated measures ANOVA test, in case the normality of the distributions is compromised. The very low p-value returned by the Friedman test confirms our conclusions from the Repeated Measures ANOVA model.

To test whether the larger data set replicates the results of the pairwise t-tests for independent observations in showing that each mask is different from every other mask and to measure the magnitude of this difference, we conduct the non-parametric, pairwise Games-Howell test. Table II shows these results. The low p-values confirm that the differences in mask performance are statistically significant throughout the data set. The difference between means column shows the magnitude of the disparity in efficacy between masks, by comparing the concentration amount that mask A filters to the concentration amount that mask B filters. For instance, the difference in concentration between cloth masks and heavy starch masks is almost equivalent to the difference between cloth masks and surgical masks, which explains that heavy starch masks and surgical masks are almost equally efficient.

**TABLE II.**
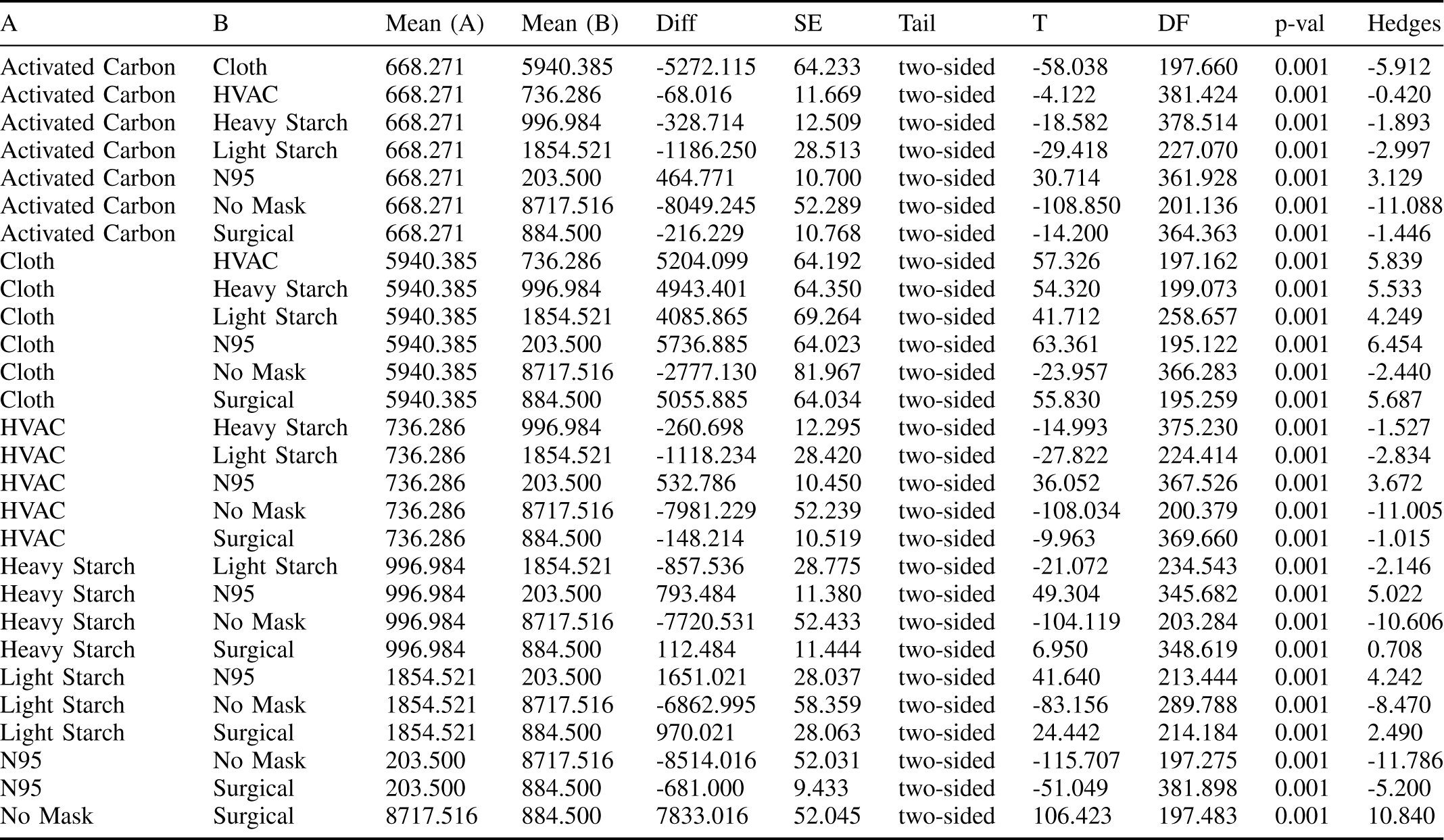
Pairwise Games-Howell test results. The results all have p-values much less than 0.001, which the test rounds to 0.001. These results demonstrate that each type of mask is significantly different from every other type of mask in its filtration capabilities. The differences between means column shows the magnitude of this difference, by comparing the concentration amount that mask A filters to the concentration amount that mask B filters. For instance, the difference in concentration between cloth masks and heavy starch masks is almost equivalent to the difference between cloth masks and surgical masks, which explains that heavy starch masks and surgical masks are almost equally efficient.

| A | B | Mean (A) | Mean (B) | Diff | SE | Tail | T | DF | p-val | Hedges |
| --- | --- | --- | --- | --- | --- | --- | --- | --- | --- | --- |
| Activated Carbon | Cloth | 668.271 | 5940.385 | -5272.115 | 64.233 | two-sided | -58.038 | 197.660 | 0.001 | -5.912 |
| Activated Carbon | HVAC | 668.271 | 736.286 | -68.016 | 11.669 | two-sided | -4.122 | 381.424 | 0.001 | -0.420 |
| Activated Carbon | Heavy Starch | 668.271 | 996.984 | -328.714 | 12.509 | two-sided | -18.582 | 378.514 | 0.001 | -1.893 |
| Activated Carbon | Light Starch | 668.271 | 1854.521 | -1186.250 | 28.513 | two-sided | -29.418 | 227.070 | 0.001 | -2.997 |
| Activated Carbon | N95 | 668.271 | 203.500 | 464.771 | 10.700 | two-sided | 30.714 | 361.928 | 0.001 | 3.129 |
| Activated Carbon | No Mask | 668.271 | 8717.516 | -8049.245 | 52.289 | two-sided | -108.850 | 201.136 | 0.001 | -11.088 |
| Activated Carbon | Surgical | 668.271 | 884.500 | -216.229 | 10.768 | two-sided | -14.200 | 364.363 | 0.001 | -1.446 |
| Cloth | HVAC | 5940.385 | 736.286 | 5204.099 | 64.192 | two-sided | 57.326 | 197.162 | 0.001 | 5.839 |
| Cloth | Heavy Starch | 5940.385 | 996.984 | 4943.401 | 64.350 | two-sided | 54.320 | 199.073 | 0.001 | 5.533 |
| Cloth | Light Starch | 5940.385 | 1854.521 | 4085.865 | 69.264 | two-sided | 41.712 | 258.657 | 0.001 | 4.249 |
| Cloth | N95 | 5940.385 | 203.500 | 5736.885 | 64.023 | two-sided | 63.361 | 195.122 | 0.001 | 6.454 |
| Cloth | No Mask | 5940.385 | 8717.516 | -2777.130 | 81.967 | two-sided | -23.957 | 366.283 | 0.001 | -2.440 |
| Cloth | Surgical | 5940.385 | 884.500 | 5055.885 | 64.034 | two-sided | 55.830 | 195.259 | 0.001 | 5.687 |
| HVAC | Heavy Starch | 736.286 | 996.984 | -260.698 | 12.295 | two-sided | -14.993 | 375.230 | 0.001 | -1.527 |
| HVAC | Light Starch | 736.286 | 1854.521 | -1118.234 | 28.420 | two-sided | -27.822 | 224.414 | 0.001 | -2.834 |
| HVAC | N95 | 736.286 | 203.500 | 532.786 | 10.450 | two-sided | 36.052 | 367.526 | 0.001 | 3.672 |
| HVAC | No Mask | 736.286 | 8717.516 | -7981.229 | 52.239 | two-sided | -108.034 | 200.379 | 0.001 | -11.005 |
| HVAC | Surgical | 736.286 | 884.500 | -148.214 | 10.519 | two-sided | -9.963 | 369.660 | 0.001 | -1.015 |
| Heavy Starch | Light Starch | 996.984 | 1854.521 | -857.536 | 28.775 | two-sided | -21.072 | 234.543 | 0.001 | -2.146 |
| Heavy Starch | N95 | 996.984 | 203.500 | 793.484 | 11.380 | two-sided | 49.304 | 345.682 | 0.001 | 5.022 |
| Heavy Starch | No Mask | 996.984 | 8717.516 | -7720.531 | 52.433 | two-sided | -104.119 | 203.284 | 0.001 | -10.606 |
| Heavy Starch | Surgical | 996.984 | 884.500 | 112.484 | 11.444 | two-sided | 6.950 | 348.619 | 0.001 | 0.708 |
| Light Starch | N95 | 1854.521 | 203.500 | 1651.021 | 28.037 | two-sided | 41.640 | 213.444 | 0.001 | 4.242 |
| Light Starch | No Mask | 1854.521 | 8717.516 | -6862.995 | 58.359 | two-sided | -83.156 | 289.788 | 0.001 | -8.470 |
| Light Starch | Surgical | 1854.521 | 884.500 | 970.021 | 28.063 | two-sided | 24.442 | 214.184 | 0.001 | 2.490 |
| N95 | No Mask | 203.500 | 8717.516 | -8514.016 | 52.031 | two-sided | -115.707 | 197.275 | 0.001 | -11.786 |
| N95 | Surgical | 203.500 | 884.500 | -681.000 | 9.433 | two-sided | -51.049 | 381.898 | 0.001 | -5.200 |
| No Mask | Surgical | 8717.516 | 884.500 | 7833.016 | 52.045 | two-sided | 106.423 | 197.483 | 0.001 | 10.840 |

After both parametric and non-parametric analysis of the data, which account for any assumptions, the results confirm that the mask types come with different filtration abilities. Thus, the masks’ efficacy in filtering PM2.5 takes the following order, from most effective to least effective: N95, activated carbon, HVAC filter, surgical, heavy starch, light starch, and regular cloth mask.

## V.Conclusions

Evaluating the efficacy of some popular and some novel low-cost materials in boosting filtration characteristics of masks can help equip communities to make and donate better protective masks with little to no additional cost (and even reducing the cost in some cases). In rural and impoverished areas where COVID-19 breakouts are occurring [12] where purchasing masks may not be an option, soaking a homemade cloth mask in rice starch can make a simple cloth mask much more effective. It is also crucial for people around the world to understand that cloth masks tend to offer a false sense of security and that they can take simple measures in their own homes to increase the efficacy of these masks threefold. With the highly contagious nature of the virus, each member of every community must take the necessary precautions to keep themselves and others safe.

The primary limitation of this study is that it has not been performed with real respiratory droplets or virus particles. We attempted to simulate such an environment with particles of a similar size and chemical nature. However, there may be some inaccuracy in the results due to differences between the simulation and real-world scenarios. In addition, due to limited resources, we were unable to test masks with a variety of brands for one kind of filter or cloth, so the efficacy of other types of cloth masks may vary. Other types of starch with differing concentrations of amylose and amylopectin may also minorly affect the filtration properties of the masks. The fit of the masks on the mannequin may not also not perfectly replicate the leakages created when worn by a person, because the focus of this study is primarily to compare the filtration abilities of the materials themselves.

## Data Availability

Data for these modeled experiments are available to other researchers. Please contact the authors. There is no Protected Health Information in this study.

